# Exercise misinformation in fitness content targeted at women on Instagram and TikTok: a cross-sectional content analysis of 6,982 posts

**DOI:** 10.64898/2026.11.19.26368260

**Authors:** Isabelle Schlegel, Ailsa Sandilands, Gozde Ozakinci

## Abstract

**Objectives:** To quantify exercise misinformation in female-targeted exercise content on Instagram and TikTok, determine whether content textually targeted at women carries higher rates of misinformation, and compare content from creators self-identifying as ethnic minority with other content.

**Methods:** Cross-sectional content analysis of 6,982 posts collected March to June 2026 by manual browsing, which captured on-screen and caption text, and automated collection, which captured captions only. Text was coded for claim accuracy, textual female targeting and creator characteristics, with large language model-assisted coding verified against an independent human coder. Effects were estimated by logistic regression using adjustment sets from a pre-specified causal diagram, with creator-clustered standard errors.

**Results:** Of 6,524 posts, 2,447 made an assessable exercise claim, of which 600 (24.5%, 95% CI 22.8 to 26.2) contained misinformation. Textually female-targeted content carried more than twice the odds of misinformation compared with untargeted content (OR 2.06, 95% CI 1.58 to 2.70), particularly wholly inaccurate content. The effect was OR 4.00 (2.41 to 6.66) where on-screen text was captured and 1.04 (0.73 to 1.49) where only captions were (interaction p < 0.001). Content from creators self-identifying as ethnic minority carried less misinformation (OR 0.69, 95% CI 0.51 to 0.92).

**Conclusion:** 24.5% of fitness posts making a verifiable claim contained misinformation, and content directed at women had higher rates where the full written text could be read. Posts naming a body part were most likely to be inaccurate, giving clinicians and public health communicators a teachable marker of misinformation.

*Key messages:* What is already known on this topic

- Health misinformation is common on social media, and fitness content aimed at women has been linked to harmful impacts on mood, body image and exercise behaviour.
- Existing research concentrates on YouTube or small samples, is rarely gender-specific, and contains no ethnic minority sub-analyses. What this study adds

- One in four posts making a verifiable exercise claim contained misinformation, and textually female-targeted content carried more than twice the odds of untargeted content — four times the odds where on-screen text (in addition to caption text) could be read.
- Content directed at women implicitly, by naming a body part, carried higher odds of misinformation than content explicitly addressing a female audience, and was more likely to be wholly inaccurate.
- Content from creators self-identifying as ethnic minority carried less misinformation, not more. How this study might affect research, practice or policy

- Women navigate a measurably worse exercise information environment on social media, and the misinformation directed at them is disproportionately about altering body shape rather than enhancing their capability; this framing is already linked to negative biopsychosocial effects on women.
- Given the content with the highest odds of being wholly inaccurate never addresses women directly, advice and policy aimed at explicitly gendered marketing will miss most of it; appraisal must be based on what a post claims, not on whom it addresses.
- Clinicians have a usable rule: content naming a body part and promising a shape warrants scepticism; content describing a practice and a capability warrants less. The element most likely to be false is the promised outcome, not the exercise.
- Commercial intent and self-described “coach” status are weak discriminators of misinformation and should not be regarded? as warning signs.

## Introduction

Physical activity is among the most effective interventions in preventive medicine,^1^ yet the information environment surrounding it is unregulated. Social media is now a routine source of health information,^2^ s and short-form fitness content reaches audiences at a scale no clinical service can match. Inaccurate content risks injury, disordered relationships with exercise and body image, financial loss through programmes sold on false premises, and erosion of trust in qualified sources.^4 5^ Clinicians increasingly meet patients whose beliefs were formed online,^6^ but are rarely equipped to advise how to appraise such content.

Content directed at women is of particular concern. Exposure to ‘fitspiration’ imagery affects mood, body image and exercise behaviour,^7^ and such content often frames exercise as a means of altering appearance rather than improving capability, a framing that invites claims exercise science does not support. Several female-targeted areas are known to be prone to misrepresentation, including sex differences in training response, menstrual cycle phase and localised fat reduction.^8^⁻^10^

There is minimal current research depicting the exercise misinformation environment on social media. A 2025 systematic review found that most work examines YouTube or small TikTok samples, is rarely gender-specific and contains no ethnic minority sub-analyses.^11^ Reliance on social media for health information is higher among ethnic minority respondents in English-speaking countries,^12 13^ yet no study has examined whether content from ethnic minority creators differs in accuracy.

Women in the United States are more likely than men to take health information from social media influencers, most markedly at ages 18–29 (57% vs 47%).^14^ This reflects need as well as preference: 84% of respondents to England’s call for evidence on women’s health reported not being listened to by healthcare professionals,^15^ and social media fills the gaps that evidence and services leave.^16^ That environment is heavily commercial: 39% of #womenshealth posts on Instagram sold a product or service, and 7% cited scientific evidence.^17^

Instagram and TikTok distribute short-form content by algorithmic recommendation rather than subscription, so posts reach users who do not follow their creators, and both have their largest audiences among adults under 35.^2^

This study quantified the prevalence of exercise misinformation in fitness content on these platforms, tested whether textually female-targeted content carries more misinformation than untargeted content, and compared content from creators self-identifying as ethnic minority with other content.

## Methods

### Design and reporting

Cross-sectional quantitative content analysis of publicly posted fitness content, reported to STROBE^18^ with items specific to social media research: search dates, personalisation control, sampling strata, retrieved versus included denominators, collection tool and data retention. Analysis was restricted to written text: caption, on-screen text where captured, hashtags and creator biography.

### Sampling and data collection

Posts were collected between March and June 2026 through three sampling routes: hashtag browsing, keyword search and the platform recommendation feed. Twenty hashtags, selected from a literature search and platform tag suggestions,^11 19^ were used across both collection arms, including four ethnic-minority-specific tags for the sub-study.

Manual collection (N = 1,200) was carried out by two researchers using dedicated research accounts that followed no creators, so the recommendation feed reflected the search terms rather than a browsing history; this arm recorded on-screen text. Automated collection (N = 5,782) used a commercial scraping tool (Apify) used in peer-reviewed health research^20 21^ and could not capture on-screen text.

On-screen text carries 66.6% of TikTok’s written content but 22.8% of Instagram’s, so platforms were compared only within a collection arm (manual or automated); combining arms would compare text availability rather than platforms.

### Coding

The primary outcome, misinformation status, had four levels: ‘not applicable’ (no assessable exercise claim), ‘no’ (all claims accurate), ‘mixed’ (accurate and inaccurate claims coexisting independently) and ‘yes’ (all claims inaccurate). An exercise prescription presented as the means to an inaccurate outcome was coded ‘yes’, since it is not an independent accurate claim.

Textual female targeting was coded as ‘explicit’ (a gendered term of address was used), ‘implicit’ (no gendered term was used, but text plainly directed at a female audience) or ‘none’. Ethnic minority status was coded only from explicit self-identification in text; inference from photographs, names or language was prohibited. Professional status required a named credential; self-description as a coach or trainer was not sufficient. Accuracy was assessed against American College of Sports Medicine guidelines^22^ and current systematic reviews,^8^⁻^10^ and content quality with the Global Quality Score.^23 24^

Coding was large language model (LLM)-assisted with human verification, under a framework reported separately.^25^ Two human coders calibrated the instrument before any AI coding took place (Gwet’s AC1 0.977 on the primary outcome). The LLM then coded the full corpus and AI coding was verified against an independent human coder blinded to the AI codes (AC1 0.847 in a representative sample, N = 150; 0.868 in an outcome-enriched sample, N = 50). AC1 was pre-specified because the outcome is heavily skewed, where Cohen’s kappa understates agreement.^26^ Variables below the pre-set threshold of 0.80 are reported as exploratory. Female targeting reached AC1 0.817 at three levels and 0.911 (95% CI 0.857 to 0.966) for any versus none; the implicit–explicit split is not reliably measured among minority-identifying creators.

### Statistical analysis

Tests were two-sided (α 0.05) and proportions use Wilson score intervals.^27^ A causal diagram specified in DAGitty^28^ before models were finalised gave each exposure its own minimal sufficient adjustment set, so every reported effect is a total effect;^29^ for female targeting the set is {ethnic minority self-identification, sampling route} (all sets in supplementary table S11). The three-level outcome was analysed by multinomial logistic regression. Because 20.3% of claim-bearing posts came from creators with more than one post, headline effects use standard errors clustered on creator. Analyses were conducted in jamovi 2.7.37^30^ and independently reproduced in Python; figures were produced in R (version 4.6.1)^31^ using ggplot2.^32^

### Patient and public involvement

No patients or members of the public were involved in the design, conduct, reporting or dissemination of this study.

### Equity, diversity and inclusion

The study addresses a gap in gender-specific and ethnicity-specific evidence and includes a pre-planned sub-study of content from creators self-identifying as ethnic minority. The author team is all-female and spans career stages from early career to senior roles. No member of the communities concerned was involved in study design, which is identified as a limitation.

### Ethics

The University of Stirling ethics panel confirmed the study as exempt from full ethics review on 30 January 2026 (reference EC 2026 26091 20102) and approved a change request on 13 November 2026 (EC 2026 26091 22574). No human participants were recruited, and data were processed under Articles 6(1)(e) and 9(2)(j) of the UK General Data Protection Regulation. Posts are not quoted verbatim.

## Results

### Sample

Of 6,982 posts retrieved, 458 were excluded as non-English, leaving 6,524 (Instagram N = 3,867, 59.3%; TikTok N = 2,657, 40.7%; automated N = 5,370, 82.3%; manual N = 1,154, 17.7%). Of these, 2,447 (37.5%) made at least one assessable exercise claim and form the denominator for all misinformation analyses (Figure 1).

**Figure 1.**
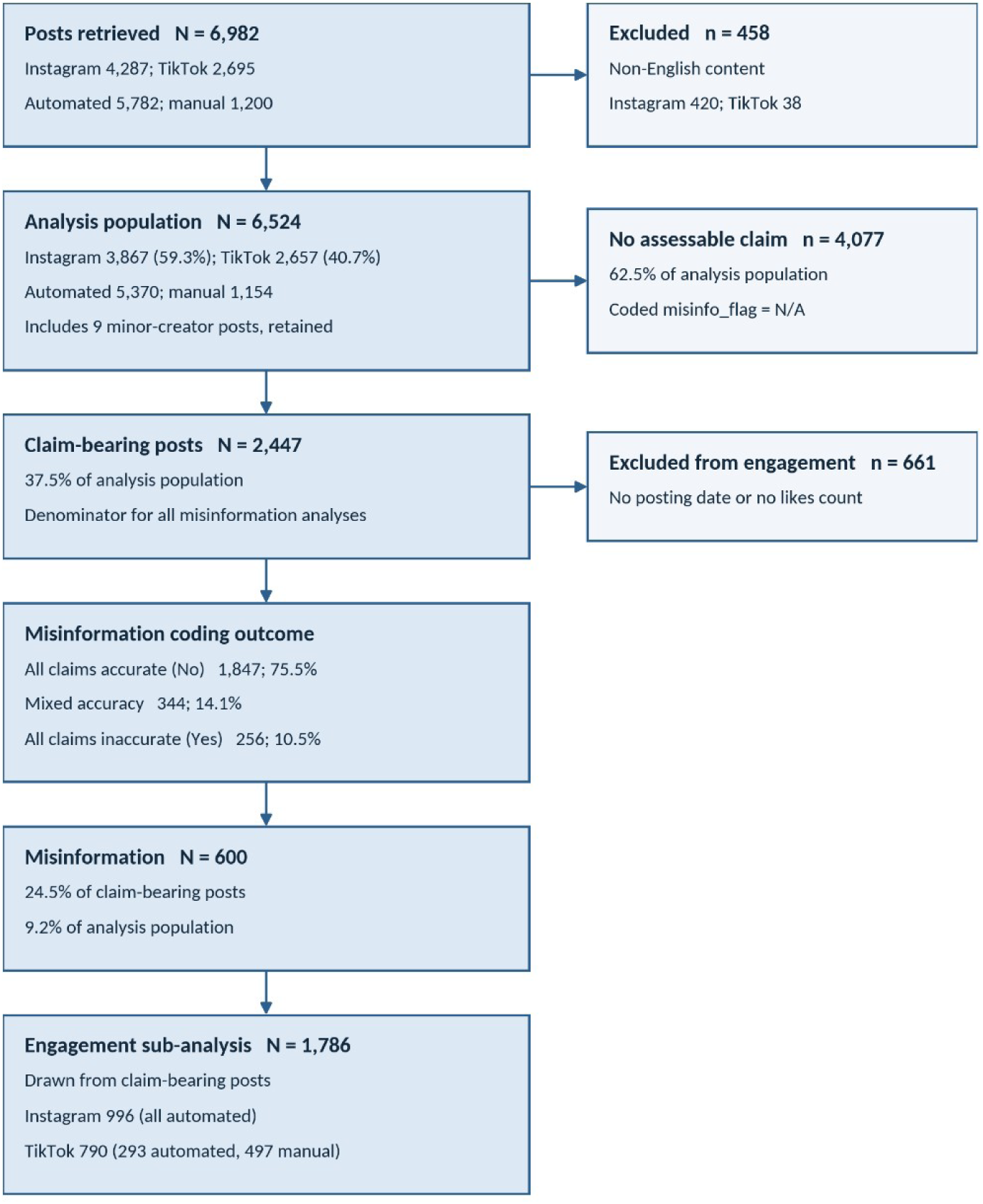
Flow of posts from retrieval to analysis, with exclusions and analytic subsets.

### Prevalence

Misinformation was present in 600 posts: 24.5% of claim-bearing posts (95% CI 22.8 to 26.2) and 9.2% of the analysis population (95% CI 8.5 to 9.9). Of these, 256 (42.7%) contained no accurate claim and 344 (57.3%) combined accurate and inaccurate claims. Both figures are lower bounds. On verification, the human coder identified misinformation more frequently than the AI coder, so residual measurement error runs toward under-detection.

### Textual female targeting

Content textually targeted at women carried more than double the odds of misinformation compared to untargeted content (OR 2.06, 95% CI 1.58 to 2.70, p < 0.001).

Explicitly female-targeted posts (N = 1,803) carried OR 1.95 (95% CI 1.12 to 3.41) and implicitly targeted posts (N = 142) OR 3.58 (95% CI 2.08 to 6.16) relative to untargeted posts (N = 502); implicit targeting carried higher odds than explicit (OR 1.72, 95% CI 1.19 to 2.49, p = 0.004).

The association sat in wholly inaccurate content. In the multinomial model, implicit targeting carried OR 9.20 (95% CI 4.62 to 18.28) for a wholly inaccurate post and no detectable association with mixed-accuracy content (OR 1.48, 95% CI 0.84 to 2.62).

The estimate for all posts combined conceals a difference between collection arms (Figure 2; Supplementary Table S12). Targeted content carried OR 4.00 (95% CI 2.41 to 6.66, N = 1,033) where caption and on-screen text were both available, and no detectable association where captions alone were (OR 1.04, 95% CI 0.73 to 1.49, N = 1,414; interaction OR 3.66, 95% CI 1.94 to 6.92, p < 0.001). In the manual arm, 32.2% of misinformation in targeted posts (114 of 354) appeared only in on-screen text. Untargeted content behaved almost identically in the two arms (13.8% vs 15.3%), which argues against manual retrieval having favoured posts containing misinformation. Implicit targeting retained its association in the caption-only arm (OR 2.44, 95% CI 1.12 to 5.31).

**Figure 2.**
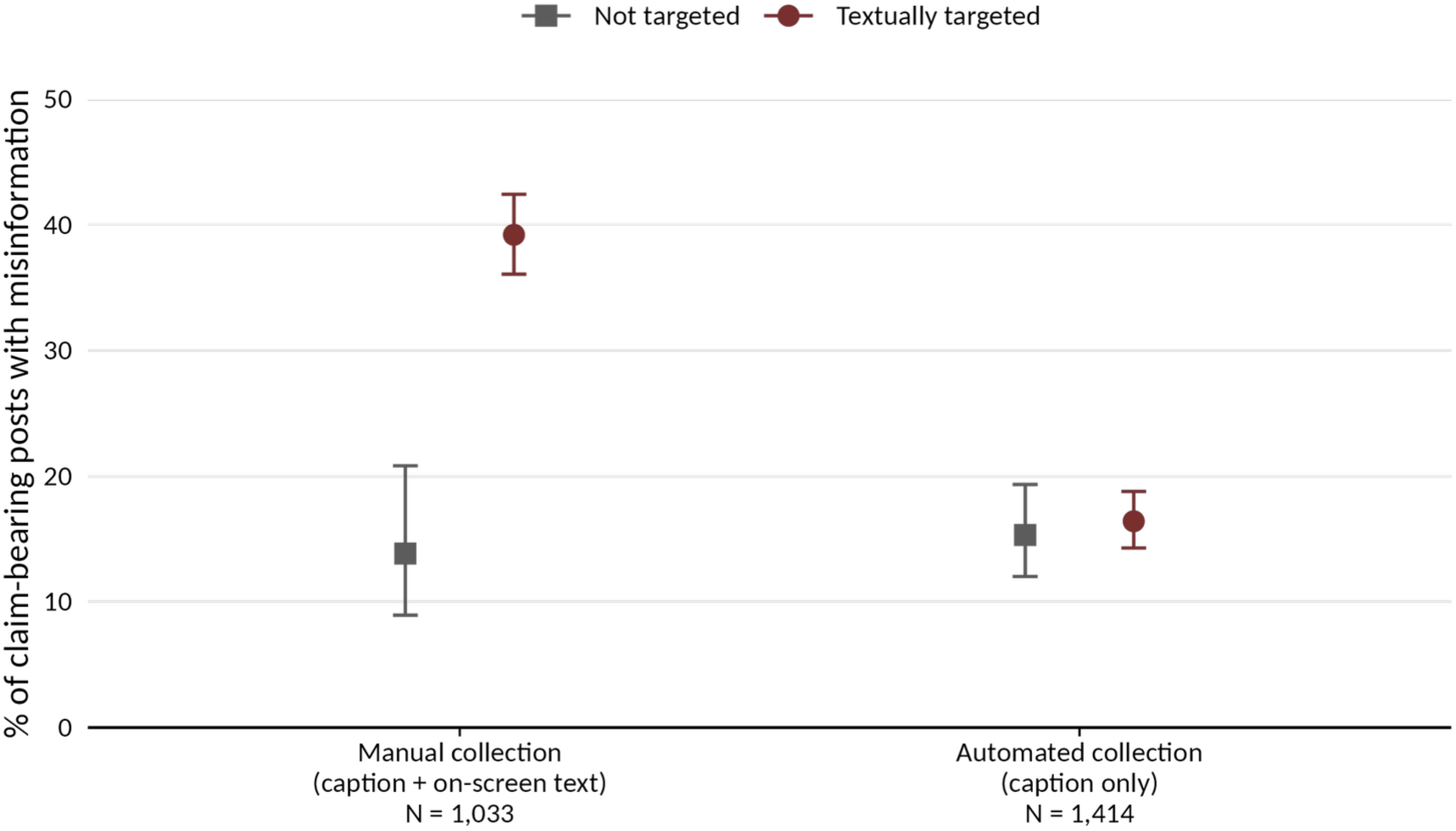
Misinformation by textual female targeting, shown separately for each collection arm, with 95% Wilson confidence intervals. The association is present where on-screen text was captured and absent where it was not.

Post hoc, 45.1% of implicitly targeted posts named the glutes versus 6.4% of untargeted posts, and of seven body regions only the glutes and waist discriminated for misinformation. Aesthetic vocabulary such as ‘tone’ and ‘sculpt’ marked targeted content generally but did not distinguish implicit from explicit.

### Platform

Within manual collection, TikTok carried higher odds than Instagram (OR 1.59, 95% CI 1.34 to 1.88). Within automated collection the difference was not significant (OR 1.23, 95% CI 0.88 to 1.72), a comparison biased against TikTok because caption-only coding removes more of its written content. TikTok scored lower on content quality in both arms (automated OR 0.60, 95% CI 0.47 to 0.76; manual OR 0.69, 95% CI 0.55 to 0.86).

### Other exposures

Total effects for the remaining exposures, each from a separate model, are shown in Figure 3. Keyword sampling carried higher odds than hashtag browsing (OR 1.76, 95% CI 1.35 to 2.29) as did the recommendation feed (OR 1.50, 95% CI 1.23 to 1.84). Monetised content carried OR 1.36 (95% CI 1.11 to 1.65, p = 0.003).

**Figure 3.**
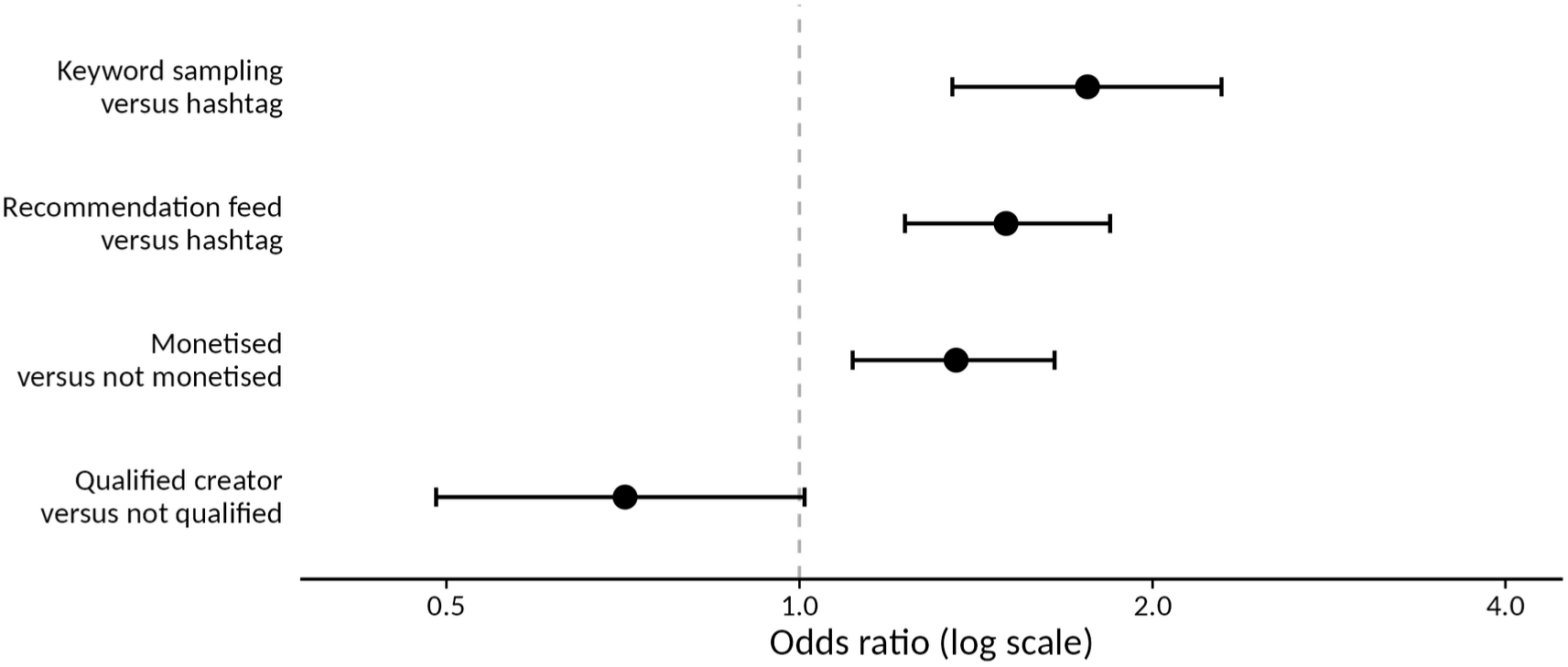
Total effects on misinformation among claim-bearing posts, with 95% confidence intervals. Each estimate derives from a separate model using its own adjustment set.

Content from qualified creators showed lower odds of misinformation, not significant once clustering standard errors at the creator level was accounted for (OR 0.71, 95% CI 0.49 to 1.01, p = 0.059; underpowered). Qualified creators scored higher on content quality (ordinal OR 1.74 to 2.19). Of 4,018 distinct creators, 19.3% described themselves as a coach or trainer, while only 6.5% named a certification and 3.2% a degree.

### Ethnic minority creators

Among claim-bearing posts from creators explicitly self-identifying as ethnic minority (N = 466), misinformation prevalence was 17.4% versus 26.2% from those not identifying as ethnic minority? (OR 0.69, 95% CI 0.51 to 0.92). Among misinformation posts, minority creators used outcome-promising language (45.7% vs 66.2%) and commercial framing (29.6% vs 46.9%) less often, and fewer rhetorical features per post (mean 1.54 vs 1.91). Implicit targeting rarely appeared in their content (Table 1).

**Table 1.**
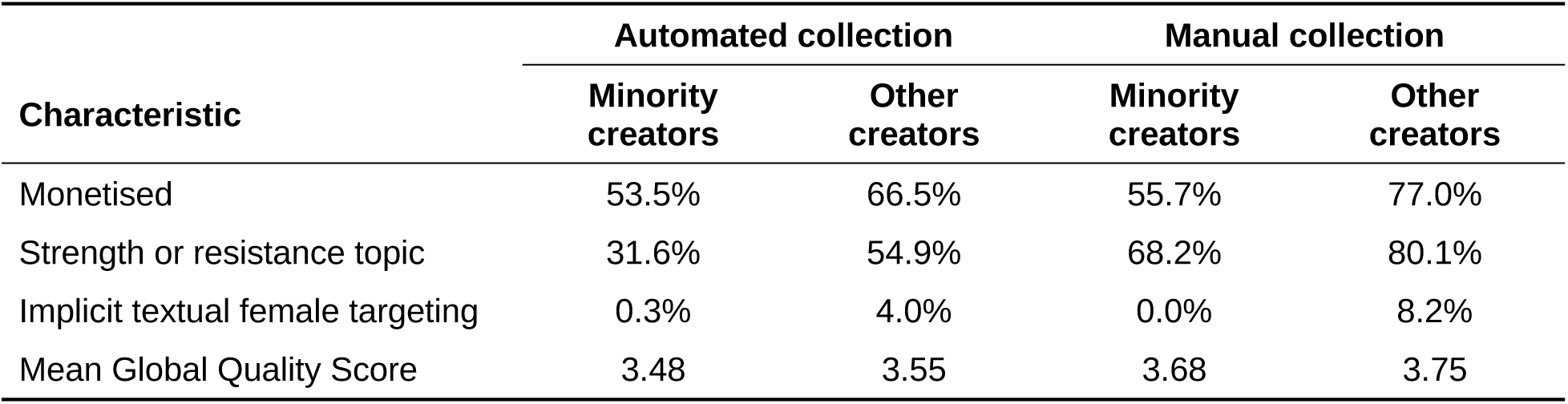
Accurate content from ethnic minority creators versus all other creators, stratified by collection method. Exercise topic (AC1 0.782) and the implicit targeting row are exploratory: the implicit–explicit split is not reliably measured among minority creators.

| Characteristic | Automated collection |  | Manual collection |  |
|---|---|---|---|---|
|  | Minority creators | Other creators | Minority creators | Other creators |
| Monetised | 53.5% | 66.5% | 55.7% | 77.0% |
| Strength or resistance topic | 31.6% | 54.9% | 68.2% | 80.1% |
| Implicit textual female targeting | 0.3% | 4.0% | 0.0% | 8.2% |
| Mean Global Quality Score | 3.48 | 3.55 | 3.68 | 3.75 |

### Engagement and lexical markers

On TikTok within manual collection, misinformation prevalence rose across tertiles of likes per day from 29.8% to 39.4% to 47.8% (p = 0.014); the most liked posts had higher rates of misinformation. The gradient was absent in the automated TikTok arm, consistent with the missing on-screen text, and on Instagram; post age did not account for this (Spearman ρ = −0.024, p = 0.499).

In caption text, ‘hourglass’ appeared 37 times more often in misinformation than in accurate content, ‘waist’ 5.7 times, ‘sculpted’ 4.6 times and ‘toned’ 3.6 times. Accurate content was characterised by capability language (‘capable’, ‘stronger’), process language (‘habits’, ‘journey’) and, most distinctively, cautious language: ‘sometimes’ appeared 8 times more often.

Hashtags carried the same signal (Table 2). Against a baseline of 24.5%, the highest rates were under body-part tags (#coreworkout 46.8%, #glutegrowth 45.9%) and the lowest under strength-identity tags (#womenwholift 6.7%, #fitnesscoach 4.3%), approximately a fourfold difference.

**Table 2.**
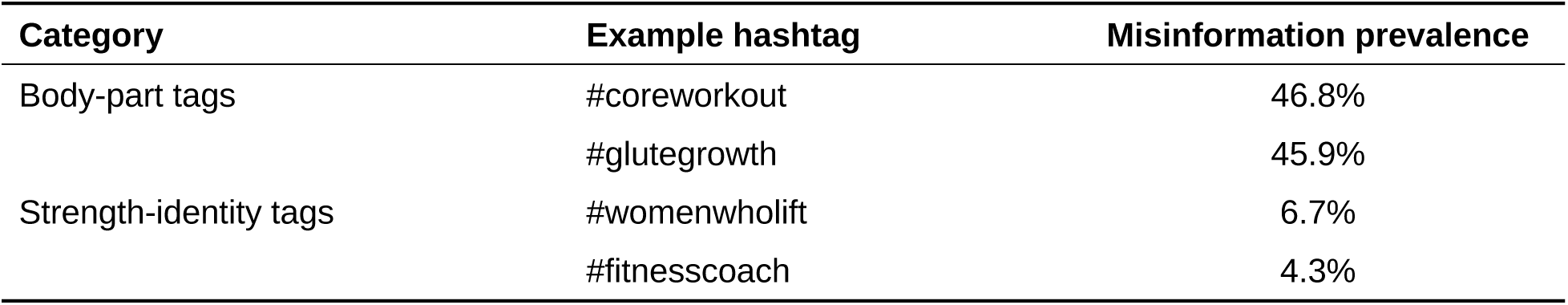
Misinformation prevalence by hashtag category, against a baseline of 24.5% among all claim-bearing posts.

## Discussion

Approximately one in four fitness posts making a verifiable exercise claim contained misinformation, and content textually targeted at women carried more than double the odds of untargeted content, rising to four times where both the caption and the on-screen text could be read. Risk of misinformation? was elevated whether targeting was explicit or implicit, which is the finding with the clearest implications for practice.

Content implicitly targeting women (for example, ‘hourglass’ body shapes) but not explicitly using female nouns or pronouns, carried higher odds of misinformation than posts using explicit female language (for example, ‘she’ or ‘girls’). Additionally, these posts were almost entirely wholly inaccurate. Consistent with the persuasion knowledge model,^35^ explicit female address may be recognised as marketing by content consumers, thereby prompting scepticism, whereas naming a body part presents as neutral instruction, hence creators may prefer to use this angle to gain engagement. However, this interpretation is exploratory, as the implicit–explicit split fell below the reliability threshold. These posts tend to be wholly inaccurate because of how they are built: a defensible routine sold as the means to an outcome that the training cannot deliver. The exercise advice is often reasonable; the promised outcome is not.

Content from creators self-identifying as ethnic minority carried less misinformation and was less commercially framed. This challenges a common assumption. Ethnic minority respondents may rely more on social media for health information,^12 13^ but that evidence describes audiences, not the accuracy of what creators post. The finding describes creators who disclosed an identity in text, not ethnic minority creators generally, and requires replication, ideally with ethnic minority women involved in study design.

### Clinical implications

The clinically important feature of this data is not the overall prevalence but its distribution. Women seeking exercise guidance on these platforms meet a materially worse information environment than the fitness audience as a whole: more than twice the odds of misinformation in the content addressed to them, and a misinformation profile dominated by misleading aesthetic claims, which formed the largest topic (37.3%) and were the most often wholly inaccurate (76%). What is disproportionately offered to women is therefore not only more often wrong, but wrong in a particular way, reframing exercise as a means of altering body shape rather than improving capability — the framing already associated with harms to mood, body image and exercise behaviour.^7^ An unregulated information environment of this shape is a plausible mechanism by which existing inequities in women’s engagement with physical activity are reproduced rather than reduced, which makes it a public health problem as well as a clinical one.

For sport and exercise medicine and primary care, two consequences follow. First, a belief formed from online fitness content warrants closer questioning when the content was directed at women, and the element most likely to be false is the promised outcome rather than the exercise itself. Interrogating the promise rather than the programme is both more accurate and less dismissive of a patient who has found an activity they will sustain, which matters when the clinical goal is long-term adherence. Second, and less intuitively, guidance organised around audience cannot work. The content carrying the highest odds of being wholly inaccurate does not explicitly textually address women at all; it names a body part. Advice to patients, public health campaigns and platform policy aimed at explicitly gendered marketing will therefore miss most of the content they are intended to catch. Appraisal must be based on what a post claims, not on whom it appears to address.

The most useful single output is a rule that does exactly that: content naming a body part and promising a specific shape warrants scepticism, whereas content describing a practice and what a person will be able to do warrants less. The hashtag finding carries the same signal before a reader engages with a post and lends itself to public-facing digital health literacy formats.

Our findings show that two commonly suspected indicators are unreliable. Commercial intent is a weak discriminator: 67% of accurate posts were also monetised, so advising patients to distrust creators who sell things would exclude much useful content. Self-description is similarly unreliable: 19.3% of creators called themselves a coach or trainer while only 6.5% named a certification. A named qualification is a better signal forthan the word ‘coach’, but not a guarantee. Clinicians who post occupy a parasocial role, followed as relatable personalities as well as experts,^36 37^ which extends their reach and their responsibility: what sport and exercise medicine clinicians post is itself part of this information environment.

### Limitations

Female targeting was coded from written text alone, and this is the principal limitation. Much of a fitness post is carried visually — who is shown exercising, what they wear, which body regions the camera emphasises — so posts directed at women only through those channels were coded untargeted. If they resemble the textually targeted groups, reported effects are understated.

The targeting association is established where the full written text was read and is not detectable from captions alone. Caption-only coding misses the third of targeted misinformation that appears only on screen, which explains part of the attenuation; whether it explains all of it cannot be established, and manual pre-screening for relevance may have contributed. The manual estimate is therefore treated as the better-measured one, and the caption-only null is not read as evidence of no effect.

Data collection method (manual versus automated) was adjusted for in every targeting model. Five of eight conditional variables and exercise topic fell below the reliability threshold, so those analyses are exploratory. The sampling frame over-represents female-targeted content; this does not bias the odds ratio, but the proportions do not estimate how much fitness content targets women. Prevalence varied from 21.7% to 32.8% by retrieval route alone, so cross-study comparison requires matched sampling. The qualification comparison had about 60% power for the observed difference. Only English-language content was analysed; 13% of claim-bearing posts carry no creator identifier, limiting clustering in that subset.

## Conclusion

Exercise misinformation is present in approximately one in four fitness posts making a verifiable claim and is substantially more common in content textually targeted at women, most of all where a post names a body part rather than an audience. Content from creators self-identifying as ethnic minority carried less misinformation, not more. Advice and policy aimed at explicitly gendered marketing will not reach the riskiest content; markers based on what a post claims will. Because women turn to social media partly where clinical care has not served them, the accuracy of what they meet there is a health equity issue. Sport and exercise medicine clinicians can act by asking patients where their information comes from, by teaching them to appraise the claim rather than the audience, and through what they themselves post.

## Supplementary material (submitted separately)

S1 full inter-rater reliability by variable; S2 misinformation topic distribution; S3 rhetorical feature frequencies; S4 lexical markers; S5 hashtag prevalence; S6 exploratory analyses of menstrual cycle, localised fat reduction and sex-difference content; S7 comparison of collection methods and sampling routes; S8 notes on the ethnic minority creator comparison (table 1); S9 analysis population and exclusions; S10 deposited materials; S11 minimal sufficient adjustment sets; S12 female targeting by collection arm. STROBE checklist uploaded separately as “Research Checklist”.

## Declarations

### Contributors

IS conceived the study, wrote the coding instrument, conducted data collection and coding, performed the analysis and drafted the manuscript. AS contributed to data collection and served as second and verifying coder. GO supervised the study and revised the manuscript. IS is the guarantor and accepts full responsibility for the finished work, had access to the data and controlled the decision to publish.

### Use of artificial intelligence

Coding of the corpus was carried out with the assistance of a large language model (Claude Opus 5), used as the primary coder under human verification within the framework described in the Methods and reported in full separately.^25^ The coding instrument was written by a human author before any LLM-assisted coding, and all AI coding was verified against an independent human coder. No AI tool is listed as an author, and the authors accept full responsibility for the accuracy and integrity of all content. Claude (Anthropic) was also used to assist with editing the manuscript for length and clarity and with preparing the figure code; the authors reviewed and approved all content.

### Funding

This work was supported by the British Association of Sport and Exercise Medicine (BASEM) Dr Vivien Lane Scholarship, jointly funded by BASEM and the Dr Vivien Lane Foundation. The funder had no role in the design, conduct, analysis or reporting of the study.

### Competing interests

None declared.

### Patient and public involvement

Patients and/or the public were not involved in the design, conduct, reporting or dissemination plans of this research.

### Patient consent for publication

Not applicable.

### Ethics approval

Reviewed by the University of Stirling ethics panel and confirmed as exempt from full ethics review on 30 January 2026 (EC 2026 26091 20102); a change request was approved on 13 November 2026 (EC 2026 26091 22574). No human participants were recruited.

### Data availability statement

Data is available in a public, open access repository. The coded dataset, analysis script, machine-readable codebook, causal model and reliability implementation are deposited at Zenodo (https://doi.org/10.5281/zenodo.22867157). The deposited dataset is anonymised for public release: it contains no raw platform text, no creator handles, and no exact dates or engagement counts.

### Provenance and peer review

Not commissioned; externally peer reviewed.

## Notes

### Competing Interest Statement

The authors have declared no competing interest.

### Author Declarations

Data were publicly available Instagram and TikTok posts, collected between March and June 2026 by manual searching and by an automated scraper (Apify). No participants were recruited or contacted. Post-level links and creator handles are not provided, to protect creator privacy. The University of Stirling ethics panel confirmed the study as exempt from full ethics review (EC 2026 26091 20102, 30 January 2026), and a change request was approved on 13 November 2026 (EC 2026 26091 22574). The anonymised dataset is at https://doi.org/10.5281/zenodo.22867157.

