## Supplemental Figures for "Exercise misinformation in fitness content targeted at women on Instagram and TikTok: a cross-sectional content analysis of 6,982 posts"

### Supplementary Table S1. Inter-rater reliability (Gwet's AC1) by variable and comparison

| Variable | Comparison | N | AC1 | 95% CI | Meets 0.80 |
|---|---|---|---|---|---|
| **Instrument calibration (human vs human)** | | | | | |
| Misinformation status | Pilot, before rules revised[a] | 100 | 0.456 | – | Superseded |
| Misinformation status | After rule revision | 57 | 0.977 | 0.931–1.000 | Yes |
| **AI pilot, initial prompt[b]** | | | | | |
| Misinformation status | AI vs IS | 57 | 0.772 | 0.634–0.909 | Superseded |
| Misinformation status | AI vs AS | 57 | 0.795 | 0.665–0.925 | Superseded |
| **Verification: primary outcome** | | | | | |
| Misinformation status | AI vs human, representative sample (Set A) | 150 | 0.847 | 0.781–0.914 | Yes |
| Misinformation status | AI vs human, outcome-enriched sample (Set B)[c] | 50 | 0.868 | 0.756–0.980 | Yes |
| **Verification: variables coded for every post (Set A)** | | | | | |
| Female targeting (3 levels) | AI vs human | 150 | 0.817 | 0.745–0.889 | Yes |
| Ethnic minority self-identification | AI vs human | 150 | 0.867 | 0.802–0.932 | Yes |
| Professional status | AI vs human | 150 | 0.825 | 0.754–0.897 | Yes |
| Monetisation | AI vs human | 150 | 0.853 | 0.785–0.922 | Yes |
| Global Quality Score (weighted AC2) | AI vs human | 46 | 0.923 | 0.886–0.960 | Yes |
| Exercise topic (5 levels) | AI vs human | 150 | 0.782 | 0.709–0.855 | No |
| **Verification: conditional variables (Sets A and B)[d]** | | | | | |
| Misuse of scientific reference | AI vs human | 57 | 0.960 | 0.903–1.000 | Yes |
| Pseudo-scientific language | AI vs human | 57 | 0.904 | 0.809–1.000 | Yes |
| Quick fix | AI vs human | 57 | 0.870 | 0.762–0.978 | Yes |
| Number of misinformation topics | AI vs human | 57 | 0.613 | 0.448–0.779 | No |

| Variable | Comparison | N | AC1 | 95% CI | Meets 0.80 |
|---|---|---|---|---|---|
| Misinformation topic (6 levels) | AI vs human | 57 | 0.609 | 0.464–0.754 | No |
| Absolute claim | AI vs human | 57 | 0.479 | 0.250–0.709 | No |
| Commercial prompt | AI vs human | 57 | 0.404 | 0.167–0.642 | No |
| Guaranteed results | AI vs human | 57 | 0.378 | 0.135–0.621 | No |
| **Post hoc contrasts and secondary estimates** | | | | | |
| Misinformation topic, aesthetic vs other[e] | AI vs human | 57 | 0.803 | 0.650–0.956 | Yes |
| Female targeting (3 levels)[f] | AI vs human, Sets A and B | 200 | 0.847 | 0.786–0.909 | Yes |
| Female targeting, any vs none[ef] | AI vs human, Sets A and B | 200 | 0.911 | 0.857–0.966 | Yes |
| **Subgroups by creator ethnicity (Sets A and B)[g]** | | | | | |
| Misinformation status, minority creators | AI vs human | 65 | 0.848 | 0.748–0.949 | Yes |
| Misinformation status, other creators | AI vs human | 126 | 0.833 | 0.756–0.910 | Yes |
| Female targeting (3 levels), minority creators | AI vs human | 65 | 0.744 | 0.609–0.879 | No |
| Female targeting (3 levels), other creators | AI vs human | 126 | 0.902 | 0.839–0.965 | Yes |
| Female targeting, any vs none, minority creators | AI vs human | 65 | 0.889 | 0.780–0.997 | Yes |

IS, lead author; AS, second human coder and verifier. [a] Superseded: two coding rules were rewritten after this round; no confidence interval is reported. [b] Blind AI coding of the second calibration sample with the initial prompt, before the calibration examples were added; superseded. [c] Drawn from posts the AI coded as containing misinformation; cannot detect false negatives. [d] Defined only for posts coded as containing misinformation; assessed on the 57 posts both coders coded as misinformation. [e] Post hoc collapse to the contrast used in analysis. [f] Secondary estimate: the Set A estimate carries the reliability claim, and non-English posts coded N/A are counted as not targeted. [g] Minority: either coder flagged the creator as self-identifying as ethnic minority; other: neither coder did. Confidence intervals use Gwet's linearisation variance, truncated at 1.

Gwet’s AC1 was pre-specified as the primary agreement statistic because the outcome distribution is heavily skewed, conditions under which Cohen’s kappa returns low values despite high observed agreement. The threshold for reliable coding was set in advance at AC1 ≥ 0.80. Variables below threshold are reported as exploratory throughout, and analyses resting on them are labelled accordingly. Cohen’s kappa was computed alongside AC1 for every comparison and is reported in the deposited codebook.

Agreement on misinformation status did not differ by creator ethnicity (AC1 0.848 vs 0.833). Agreement on female targeting was lower for minority creators (0.744 vs 0.902) because the AI coded 8 of 11 implicitly targeted posts from minority creators as explicit; the any-versus-none distinction held (0.889).

The coding framework and its verification are reported in full separately.

### Supplementary Table S2. Distribution of misinformation topics

Among 600 posts containing misinformation.

| Topic | N | % of misinformation | % wholly inaccurate | Mean GQS |
|---|---|---|---|---|
| Misleading aesthetic claims | 224 | 37.3 | 76 | 1.91 |
| Physiological or biomechanical errors | 140 | 23.3 | 19 | 2.48 |
| Misuse of training principles | 103 | 17.2 | 24 | 2.45 |
| Overgeneralised advice | 91 | 15.2 | 21 | 2.55 |
| Hormonal pseudoscience | 31 | 5.2 | 48 | 2.26 |
| Injury-prevention myths | 11 | 1.8 | 0 | 2.64 |

GQS, Global Quality Score.

**Two qualifications apply.** The six-level topic variable reached AC1 0.609, below the pre-set threshold, and is reported as exploratory; only the binary distinction between misleading aesthetic claims and all other topics reached acceptable reliability (AC1 0.803), and that collapse was identified post hoc. Topics are also not mutually exclusive: the coding records the predominant topic only, and 57.0% of misinformation posts fell under more than one topic (mean 1.62 topics per post). This distribution should not be read as a measured prevalence of topics.

### Supplementary Table S3. Rhetorical features among misinformation posts

Among 600 posts containing misinformation; mean 1.85 features per post.

| Feature | N | % of 600 | AC1 | Status |
|---|---|---|---|---|
| Guaranteed results | 380 | 63.3 | 0.378 | Exploratory |
| Commercial prompt | 267 | 44.5 | 0.404 | Exploratory |
| Absolute claim | 233 | 38.8 | 0.479 | Exploratory |
| Pseudo-scientific language | 141 | 23.5 | 0.904 | Reliable |
| Quick fix | 72 | 12.0 | 0.870 | Reliable |
| Misuse of scientific reference | 20 | 3.3 | 0.960 | Reliable |

Four features distinguished wholly inaccurate from mixed-accuracy posts at Bonferroni-corrected significance. Directionally, wholly inaccurate posts led on guaranteed results and quick fixes, whereas mixed-accuracy posts showed more absolute claims and pseudo-scientific language. Guaranteed results and absolute claims fall below the reliability threshold, so this pattern is reported with that limitation; the two reliably coded features alone show the same split.

### Supplementary Table S4. Lexical markers

Terms appearing in at least 25 posts, compared between misinformation and accurate content at $p < 0.001$. Analysis conducted directly on caption text, since a measure derived from text avoids the reliability limitations of the coded features.

| Term | Relative frequency in misinformation captions |
|---|---|
| hourglass | 37× |
| waist | 5.7× |
| sculpted | 4.6× |
| toned | 3.6× |

Accurate content was characterised by capability language (‘capable’, ‘stronger’, ‘healthier’), process language (‘habits’, ‘journey’), and most distinctively by cautious language: ‘sometimes’ appeared 8 times more often in accurate captions than in misinformation captions.

### Supplementary Table S5. Misinformation prevalence by hashtag category

Against a baseline prevalence of 24.5% among all claim-bearing posts.

| Category | Example hashtag | Misinformation prevalence |
|---|---|---|
| Body-part tags | #coreworkout | 46.8% |
| | #glutegrowth | 45.9% |
| Strength-identity tags | #womenwholift | 6.7% |
| | #fitnesscoach | 4.3% |

Body-part hashtags carried approximately four times the misinformation of strength-identity hashtags. A full per-hashtag breakdown can be generated from the deposited dataset.

### Supplementary Table S6. Exploratory analysis of areas of known vulnerability

Three areas of exercise science identified in the introduction as prone to misrepresentation were examined by keyword search of caption text. This analysis was not pre-specified and is reported as exploratory.

| Area | N | Misinformation prevalence | Comparison | p |
|---|---|---|---|---|
| Menstrual cycle or hormonal language | 147 | 38.8% | 23.6% of other claim-bearing posts | < 0.001 |
| Localised fat reduction | 29 | 31.0% | 24.4% | 0.41 |
| Sex differences in resistance training | 105 | 29.5% | 24.3% | 0.22 |

Of posts using menstrual cycle or hormonal language, 90% were explicitly female-targeted. The coded topic of hormonal pseudoscience (N = 31) was 48% wholly inaccurate and 94% explicitly female-targeted.

Explicit references to localised fat reduction were rare. The construct is more often represented in the coding by misleading aesthetic claims, whose characteristic form is a routine directed at a body region and presented as capable of altering its shape, rather than by an explicit spot-reduction claim.

### Supplementary Table S7. Comparison of collection methods

The two-arm design permitted a comparison not originally planned.

| Measure | Automated collection | Manual collection |
|---|---|---|
| Posts collected | 5,782 | 1,200 |
| In analysis population | 5,370 | 1,154 |
| Proportion making an assessable claim | 26.3% | 89.5% |

**This comparison is confounded and is not interpretable as a characteristic of either method.** Manual browsing involved the collector selecting posts that appeared relevant at the point of collection, whereas automated collection did not. The difference therefore largely reflects pre-screening.

#### Isolating the effect of on-screen text

The effect of on-screen text capture can be isolated within the manual arm, where all posts were pre-screened equivalently.

| Platform | With on-screen text captured | Without | p |
|---|---|---|---|
| TikTok | 91.6% made a verifiable claim | 64.7% (N = 17) | < 0.001 |
| Instagram | 88.8% | 84.3% (N = 70) | 0.264 |

The asymmetry is consistent with where each platform places its written content. Among manually collected posts, on-screen text accounted for 66.6% of TikTok's written content versus 22.8% of Instagram's. Caption-only collection therefore removes a substantially greater share of assessable material from TikTok than from Instagram, which is why platforms are compared only within a collection arm in the main analysis.

**Sampling route**

| Route | Misinformation prevalence | Wholly inaccurate |
|---|---|---|
| Keyword search | 32.8% | — |
| Recommendation feed | 29.4% | 16.5% |
| Hashtag browsing | 21.7% | 9.6% |

Sampling route is confounded with collection method in this study, so these comparisons are indicative only. They are reported because the spread — 21.7% to 32.8% within a single dataset coded by a single framework — bears directly on the interpretation of prevalence estimates across the literature.

### Supplementary note S8. Ethnic minority creator comparison: notes to table 1

The differences shown in table 1 of the main text held within both collection arms and are therefore unlikely to be sampling artefacts. Two rows are exploratory: exercise topic is below the reliability threshold (AC1 0.782), and the split between implicit and explicit targeting is not reliably measured among minority creators, since the AI coded 8 of 11 implicitly targeted posts from minority creators as explicit. Explicit female targeting was higher among minority creators, but this is not reportable as a finding, since all four ethnic-minority hashtags used in collection are themselves female-targeted terms.

Misinformation topics did not differ between minority and other creators ($\chi^2 = 3.95$, df = 5, $p = 0.556$). Because the misinformation topic variable falls below the reliability threshold, this null is inconclusive rather than evidence of equivalence.

### Supplementary note S9. Analysis population and exclusions

Of 6,982 posts retrieved, 458 were excluded as non-English, leaving an analysis population of 6,524.

Nine posts carried an indication that the creator was under 18 years old. These met the inclusion criteria and were not excluded on eligibility grounds; none contained an assessable exercise claim, so all nine were coded as making no assessable claim and none contributes to any reported analysis. Eight of the nine, in which the creator stated an age under 18, are withheld from the public deposit (supplementary note S10); this lowers full-population denominators in the deposited data by eight and affects no claim-bearing analysis.

Thirteen per cent of claim-bearing posts carry no creator identifier, so clustering by creator could not be fully accounted for within that subset.

### Supplementary note S10. Deposited materials

The coded dataset, annotated analysis script, machine-readable codebook with per-variable reliability, prompt scaffold, causal model, reliability implementation and versioned study protocol with dated amendments are deposited at Zenodo (https://doi.org/10.5281/zenodo.22867157).

The deposited dataset is anonymised for public release: creator handles are replaced with sequential codes, post URLs are removed, and no re-identification key is deposited. No raw platform content is included — caption text, on-screen text, creator biographies and hashtag lists are replaced with derived indicators, and term frequencies are deposited as aggregate counts. Because exact posting dates and engagement counts could together locate a post on the platform, these are replaced with collection month, post-age band, within-platform engagement tertile and an engagement rank. Every analysis reported here can be reproduced from the deposited files, including the text-derived findings; the analysis script gives identical output on the deposited and source data. A new text analysis using different word lists cannot, and would require collecting a fresh corpus.

### Supplementary Table S11. Minimal sufficient adjustment sets

Derived in DAGitty from the causal diagram deposited as dag_model.txt. Each exposure is modelled separately against the misinformation outcome using its own set, so every reported estimate is a total effect. Where two minimal sets exist, either is sufficient; the first listed is the one used.

| Exposure | Minimal sufficient adjustment set | Equivalent alternative |
|---|---|---|
| Textual female targeting | ethnic minority self-identification, sampling route | topic, collection method |
| Platform | collection method | — |
| Creator professional status | none required | — |
| Monetisation | creator professional status | — |
| Misinformation topic | female targeting, collection method | ethnic minority, collection method, sampling route |
| Ethnic minority self-identification | none required | — |
| Sampling route | none required | — |
| Collection method | none required | — |

Engagement is a descendant of the outcome in the diagram and is therefore not treated as an exposure for misinformation; the engagement analysis is reported in the reverse direction and as exploratory.

### Supplementary Table S12. Female targeting by collection arm

Adjusted for {ethnic minority self-identification, sampling route}, with cluster-robust standard errors on creator.

| Model | N | OR | 95% CI | p |
|---|---|---|---|---|
| Any targeting vs none, all posts combined | 2,447 | 2.06 | 1.58 to 2.70 | < 0.001 |
| Any targeting vs none, caption + on-screen text | 1,033 | 4.00 | 2.41 to 6.66 | < 0.001 |
| Any targeting vs none, caption only | 1,414 | 1.04 | 0.73 to 1.49 | 0.81 |
| Targeting × collection arm interaction | 2,447 | 3.66 | 1.94 to 6.92 | < 0.001 |
| Implicit vs none, caption only | 425 | 2.44 | 1.12 to 5.31 | 0.025 |

Unadjusted percentages of claim-bearing posts containing misinformation, with Wilson 95% confidence intervals:

| Targeting | Caption + on-screen text | Caption only |
|---|---|---|
| Not targeted | 13.8% (8.9 to 20.8), n = 130 | 15.3% (12.0 to 19.3), n = 372 |
| Explicit | 38.3% (35.1 to 41.7), n = 814 | 15.7% (13.5 to 18.1), n = 989 |
| Implicit | 47.2% (37.2 to 57.5), n = 89 | 30.2% (19.5 to 43.5), n = 53 |

Sampling route does not account for the association: adding route to the model for all posts combined moves the estimate from 2.10 unadjusted to 1.99. Route composition is similar across targeting levels, and route is in the adjustment set for every targeting model reported.
